# Cross-reactive antibody responses against SARS-CoV-2 and seasonal common cold coronaviruses

**DOI:** 10.1101/2020.09.01.20182220

**Authors:** Shelley Klompus, Sigal Leviatan, Thomas Vogl, Iris N. Kalka, Anastasia Godneva, Eilat Shinar, Adina Weinberger, Eran Segal

## Abstract

Beyond SARS-CoV-2, six more coronaviruses infect humans (hCoVs), four of which cause only mild symptoms (seasonal/common cold hCoVs) ^12^. Previous exposures to seasonal hCoVs may elicit immunological memory that could benefit the course of SARS-CoV-2 infections ^3^. While cross-reactive T cells epitopes of SARS-CoV-2 and seasonal hCoVs have been reported in individuals unexposed to SARS-CoV-2 ^4-6^, potential antibody-based cross-reactivity is incompletely understood.

Here, we have probed for high resolution antibody binding against all hCoVs represented as 1,539 peptides with a phage-displayed ^7^ antigen library. We detected broad serum antibody responses against peptides of seasonal hCoVs in up to 75% of individuals. Recovered COVID-19 patients exhibited distinct antibody repertoires targeting variable SARS-CoV-2 epitopes, and could be accurately classified from unexposed individuals (AUC=0.96). Up to 50% of recovered patients also mounted antibody responses against unique epitopes of seasonal hCoV-OC43, that were not detectable in unexposed individuals.

These results indicate substantial interindividual variability and antibody cross-reactivity between hCoVs from the direction of SARS-CoV-2 infections towards seasonal hCoVs. Our accurate high throughput assay allows profiling preexisting antibody responses against seasonal hCoVs cost-effectively and could inform on their protective nature against SARS-CoV-2.

## Main text

COVID-19 (coronavirus disease 2019), caused by SARS-CoV-2 (severe acute respiratory syndrome coronavirus 2), represents an unparalleled pandemic with millions of cases worldwide. In addition to SARS-CoV-2, six more coronaviruses infect humans (hCoVs) including SARS-CoV-1 responsible for the SARS outbreak in 2003 and MERS-CoV (Middle East respiratory syndrome) ^1^. Four seasonal endemic hCoVs (OC43, HKU1, NL63, 229E) are widely circulating in the population causing only mild symptoms (common cold)^2^. Previous exposures to seasonal hCoVs may elicit immunological memory that could benefit the course of SARS-CoV-2 infections ^3^, which can range from asymptomatic to life-threatening symptoms ^8^. The exact causes underlying this heterogeneity in COVID-19 severity are incompletely understood and involve factors as age, gender, comorbidities, and preexisting immunity ^9,10^. As an important part of the adaptive immune system, it has been demonstrated that up to ca. 60% of individuals unexposed to SARS-CoV-2 show CD4+ T cell recognition of its epitopes ^4,5^ and cross-reactivity against seasonal hCoVs ^6^. Preexisting T cell or antibody responses need careful consideration in vaccine development, with recommendations to assess existing immunity in vaccine trial participants to ensure even distributions between testing groups ^3^.

Compared to studying T cell epitope recognition (which involves living cells, antigen presentation by variable MHC alleles, and rather low affinity interactions ^11^), antibody-antigen binding is robust, easily detectable, and amenable to high throughput methods (e.g. ^7,12^). Antibody tests for hCoV cross-reactivity could be valuable tools to assess preexisting immunity at large-scale and stratify vaccine trials cost effectively. Such serological testing could also inform on a possible impact of seasonal hCoVs to herd immunity against SARS-CoV-2 ^13^. Yet, cross-reactive antibody responses between SARS-CoV-2, seasonal hCoVs, and their protective potential are incompletely understood. It has been demonstrated that antibodies of COVID-19 patients cross-react against full-length spike (S) and nucleocapsid (N) proteins of SARS-CoV-1 and MERS-CoV ^14,15^. However, there are differences in the binding of protein segments, with antibodies binding the receptor-binding domain (RBD) of the SARS-CoV-2 S-protein generally failing to bind this region of SARS-CoV-1 or MERS-CoV^14,16,17^. Similarly, no cross-reactivity between antibodies against the RBD of SARS-CoV-2 and seasonal hCoVs NL63/229E was detectable ^18^. Epitope resolved antibody binding data beyond the S-protein/RBD is scarce for SARS-CoV-2, and virtually unavailable for seasonal hCoVs ^19^.

Here, we have applied a high resolution antibody assay ^7,12^ to test for binding against 1,539 peptide antigens covering all known proteins of all hCoVs. We have detected a high seroprevalence of seasonal hCoVs in up to 75% of individuals both unexposed to SARS-CoV-2 and recovered from COVID-19, variability in antibody repertoires against SARS-CoV-2, and cross-reactivity against seasonal hCoVs upon SARS-CoV-2 infection.

## Results and discussion

### Antibody repertoires against hCoVs and cross-reactivities

Antibody binding against SARS-CoV-2 is typically assessed by ELISAs against full length proteins/domains ^20,21^, by resolving crystal structures ^14,22^, or by peptide arrays ^23,24^. Pinpointing protein segments recognized by cross-reactive antibodies of all hCoVs requires high resolution and high-throughput methods. Phage immunoprecipitation sequencing (PhIP-Seq) relies on the display of synthetic oligo libraries on T7 phages ^7,12^. Thereby displayed antigens can be rationally selected allowing to probe for hundred thousands of antigens in parallel. After mixing of the phage library with serum antibodies, unbound phages are washed away after immunoprecipitation and enriched phages are detected by next generation sequencing (Fig. 1a).

**Fig. 1:**
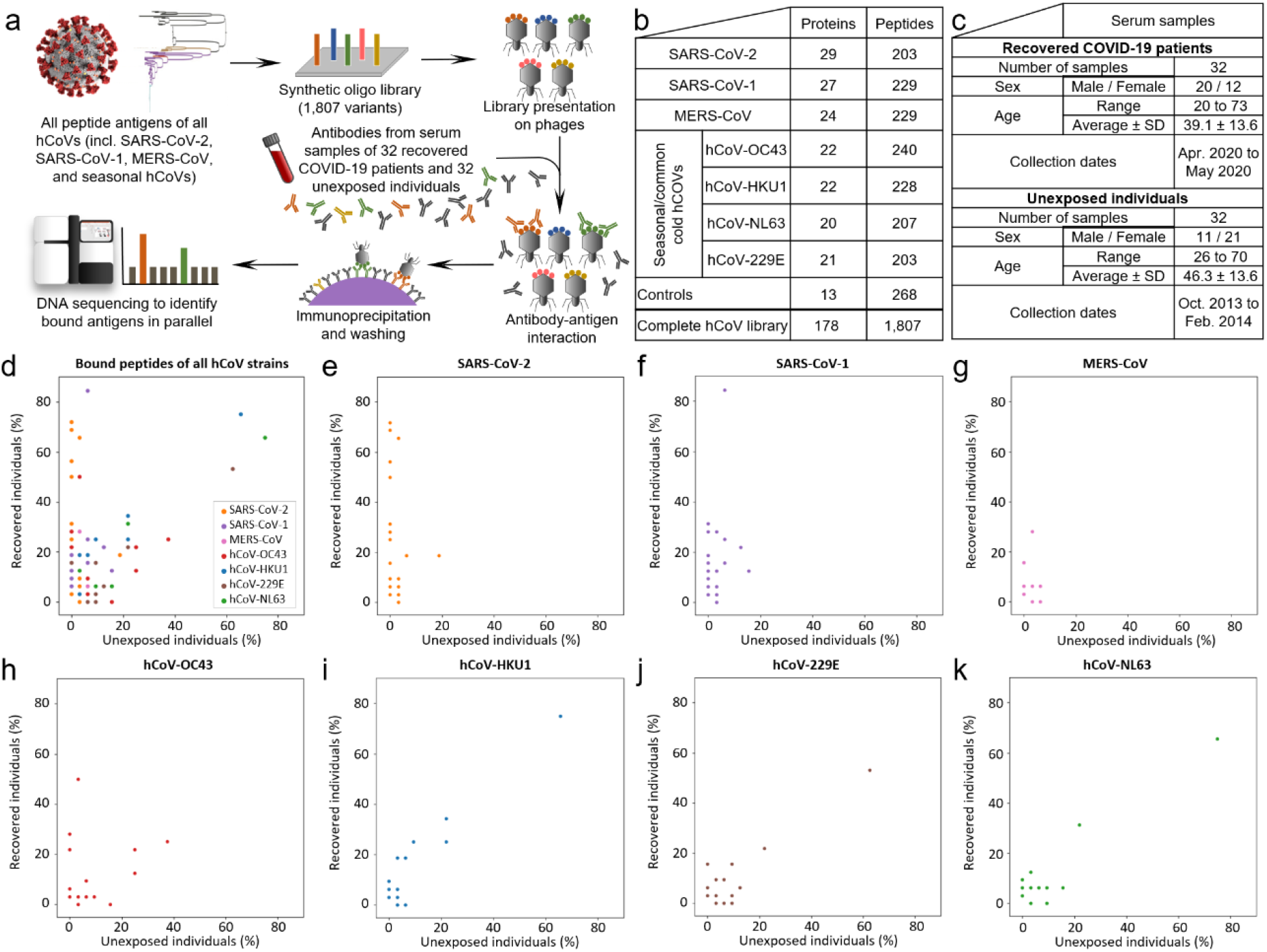
A phage displayed antigen library (a) of human Coronaviruses’ peptide antigens (b) applied to serum samples of unexposed individuals and recovered COVID-19 patients (c) detects a high serum prevalence of seasonal hCoVs, interindividual variability of antibody repertoires against SARS-CoV-2, and cross-reactive antibody responses from SARS-CoV-2 infection (d-k). The numbers of proteins per strain in panel b include polyproteins being split into 14 separate proteins. d-k, All bound antigens of the hCoV library are shown in panel d, the following panels depict binding against peptides of each hCoV strain separately. The illustration of the SARS-CoV-2 virion is reproduced from CDC PHIL #23312 released as public domain (CDC/ Alissa Eckert, MSMI; Dan Higgins, MAMS), the phylogenetic tree is reproduced form Wu et al. ^1^ [open access].

We have generated a PhIP-Seq library covering all open reading frames of hCoVs as 64 amino acid (aa) sections with 20 aa overlaps between adjacent peptides (Fig. 1b). The library also includes positive controls which confirmed detection of antibody responses against viruses previously reported to elicit population wide immunity ^12^ and negative controls, that did not show substantial binding (Table S 1).

We tested IgG antibody binding against this hCoV library with 32 serum samples of individuals unexposed to SARS-CoV-2, that had been collected in 2013/2014 ^25^ before the COVID-19 outbreak (Fig. 1c). These antibody repertoires were compared to 32 samples of recovered COVID-19 patients obtained in April and May 2020. In total we have assayed for nearly 100,000 antibody-peptide interactions (1,539 hCoV peptides in each of the 64 individuals). Employing strict Bonferroni correction, 240/1,539 peptides were enriched in at least one individual, with an average of 9 hCoV peptides significantly bound per unexposed individual and 19 hCoV peptides in recovered COVID-19 patients. Most analyses were based on antibody responses against 57 hCoV peptides from 32 different hCoV proteins, shared by more than five individuals of either group and seven peptides showing significantly different abundances between the groups (Table S 2).

Unexposed individuals showed abundant antibody responses against all seasonal hCoVs: Binding against peptides of hCoV-NL63 was detected in 75% of unexposed individuals, against hCoV-HKU1 in 66%, against hCoV-229E in 63%, and against hCoV-OC43 in 38% (Fig. 1h-k, Table S 2). The same peptides were bound at similar frequencies in recovered COVID-19 patients and originated mostly from S- or N-proteins (Table S 2). Binding of any peptide from seasonal hCoV S- or N-proteins was detectable in 78% of unexposed individuals for hCoV-NL63, 75% for hCoV-HKU1, and 63% for hCoV-229E and hCoV-OC43 with these epitope resolved results being in agreement with previous studies on the seroprevalence of seasonal hCoVs using ELISAs ^26^.

Recovered COVID-19 patients’ sera showed, as expected, an overrepresentation of several peptides of SARS-CoV-2 that completely lacked binding in unexposed individuals, with five peptides passing FDR (false discovery rate) correction for being significantly different between the two groups of individuals (Table S 2). While nearly all COVID-19 patients showed binding against at least one peptide in S- or N-proteins and some peptides being bound in up to 72% of recovered patients (Fig. 1e), no convergence of antibody responses against the same peptide were detected in all individuals. This finding differs from near universal recognition of some viral epitopes previously observed for other human viruses (*12*) and replicated with controls in this study (S 1), suggesting that the antibody response against SARS-CoV-2 can exhibit substantial inter-individual variability.

COVID-19 serum samples showed also common binding against SARS-CoV-1, indicating detection of cross-reactivity of antibodies targeting SARS-CoV-2 (Fig. 1f) (*14, 15*), most notably one SARS-CoV-1 spike peptide had significantly enriched binding in up to 84% of COVID-19 recovered individuals compared to 6% of unexposed individuals (Table S 2). A non-structural protein (NSP3) of SARS-CoV-1 was even bound in 13% of unexposed individuals and 22% of recovered, possibly owing to higher conservation of such NSPs underlying less selective pressure than S- and N-proteins mostly responsible for infectivity and targeted by neutralizing immune responses. A few other peptides were differentially enriched between the two groups, but did not pass FDR correction for significance of this difference, including a peptide of the MERS-CoV S-protein bound in 28% of COVID-19 sera and 3% (1/32) of unexposed individuals (Fig. 1g).

Strikingly, cross-reactive responses from SARS-CoV-2 also extended to the seasonal hCoVs-OC43 (Fig. 1h). One hCoV-OC43 spike peptide was bound in 50% of COVID-19 sera and 3% (1/32) of unexposed individuals, passing FDR correction for being differentially enriched between the two groups (Table S 2). Another two peptides of hCoV-OC43 spike were not bound in unexposed individuals at all, but bound in up to 28% of COVID-19 patients, neither passing FDR correction (Table S 2). Differential binding of hCoV-HKU1 was less pronounced with one peptide occurring in 19% of COVID 19 patients and 3% (1/32) of unexposed individuals (Fig. 1i) not passing FDR correction (Table S 2). We did not detect cross-reactivities against peptides from the alpha coronaviruses (hCoV-NL63 and CoV-229E), with COVID-19 patients and healthy individuals’ sera reacting at similar rates (Fig. 1j,k). Peptides eliciting cross-reactive antibody responses between SARS-CoV-2, SARS-CoV-1, and seasonal hCoVs typically originated from similar regions of S-(Fig. 2a) and N-(Fig. 2b) proteins. This data indicates that epitopes in the SARS-CoV-2 spike S2 region are frequently bound in COVID-19 patients and a target for cross-reactivity. Many recombinant SARS-CoV-2 vaccines in development focus either on the full-length spike or the RBD alone ^27^. The observed S2 reactivity could lead to differences between these designs, with responses against full-length S-protein vaccines potentially benefiting from cross-reactivities against seasonal hCoVs, which may not occur for RBD only vaccines.

**Fig. 2:**
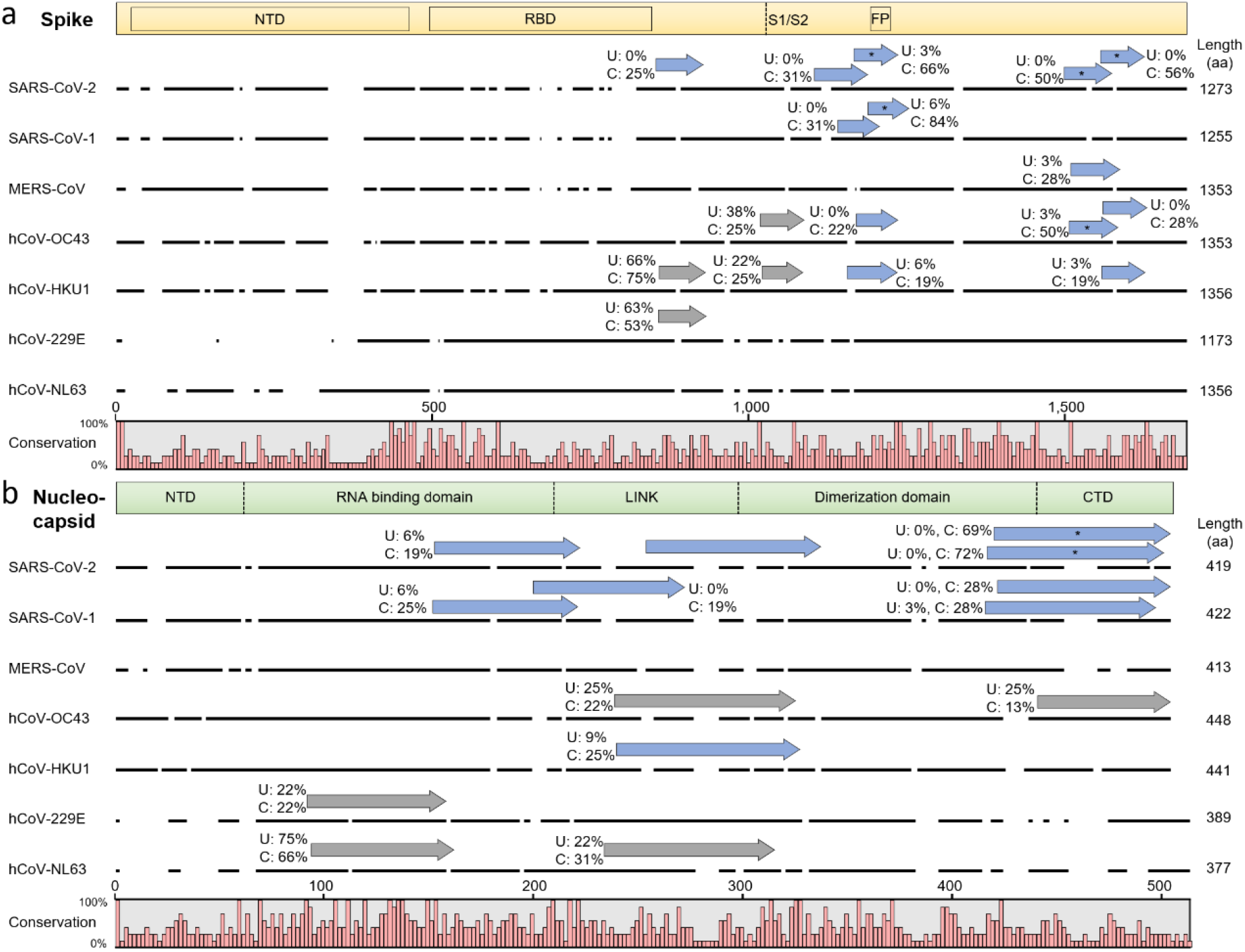
Cross-reactive and selective antibody binding of SARS-CoV-2 peptides and other hCoVs clusters in similar regions of the spike (a) and nucleocapsid (b) protein. Alignments of S- and N-proteins of all hCoVs, the dark line next to the strain identifier represents the protein sequence indicating gaps in the consensus alignment. Peptides bound in more than five unexposed (‘U’) individuals or recovered COVID-19 patients (‘C’) are shown as arrows above the corresponding protein sequence. The abundance of binding in ‘U’ and ‘C’ is indicated as percentages written next to the peptides. Grey arrows indicate similar recognition in unexposed individuals and COVID-19 patients, blue arrows indicate more than two-fold overrepresented binding in COVID-19 patients. Peptides marked with an asterisk appear at significantly different abundances (passing FDR correction) between unexposed individuals and recovered COVID-19 patients, by scoring the difference between the two distributions of the log fold changes (number of reads of bound peptides vs. baseline sequencing of phages not undergoing IPs) of the two groups (Table S 2). The domain structure on top of each panel is based on SARS-CoV-2 S-^35^/N-^36^ proteins, positions in other hCoVs shift along the alignment. Due to the different lengths of S- and N-proteins, the two panels are not drawn at the same scale.

When looking at the principal components of an analysis (PCA, Fig. 3a) performed on the log fold change (number of reads of bound peptides vs. baseline sequencing of phages not undergoing immunoprecipitation) of significantly enriched oligos, unexposed individuals clustered together while recovered COVID-19 patients’ samples showed a greater spread. This illustrates the interindividual variability in hCoV antibody responses elicited by SARS-CoV-2.

**Fig. 3:**
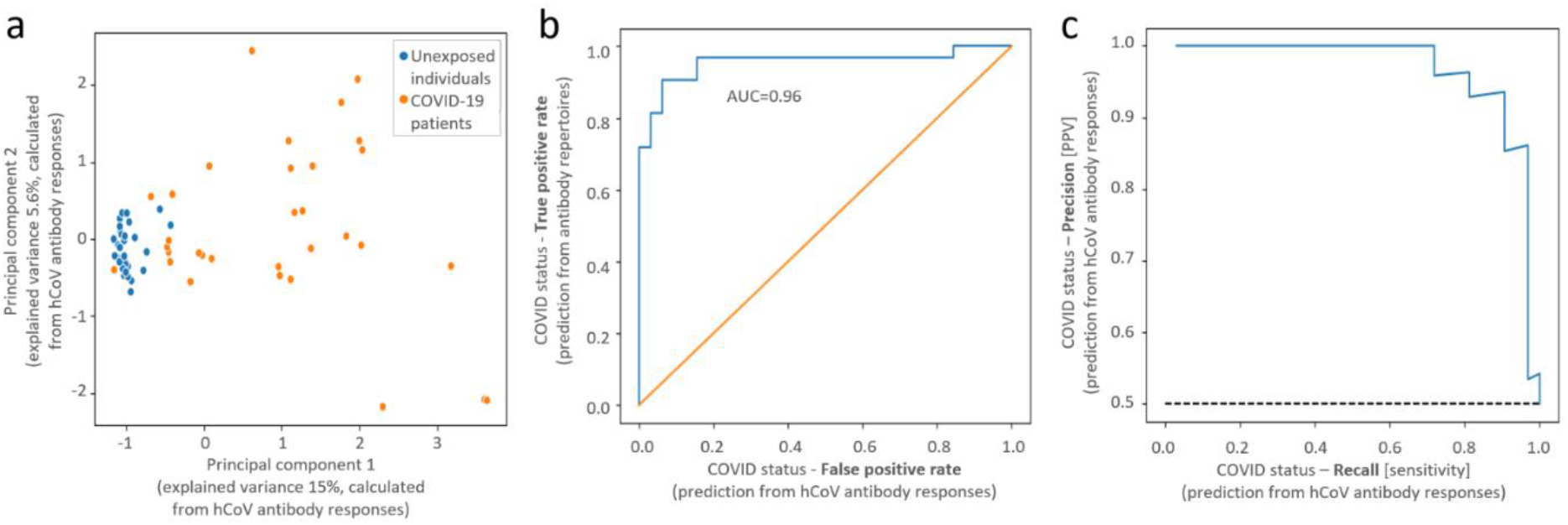
Principal component analysis (PCA) separates between hCoV antibody responses of unexposed individuals and recovered COVID-19 patients (a) and a machine learning predictor accurately identifies infected individuals (b,c). Gradient boosting decision trees (XGBoost classifier ^37^) with leave-one-out cross-validation were used for these predictions on hCoV peptides bound in >4 (5%) individuals. Area under the curve (b) and precision-recall curve (c) of predicting COVID-19 status from antibody responses against hCoV peptides (abbreviations: PPV - positive predictive value).

Overall, this data indicates that substantial cross-reactivity between hCoVs observed for T cells ^4,5^ also extends to antibody responses against seasonal hCoVs. We show that infection with SARS-CoV-2 mounts cross reactive antibodies against hCoV-OC43 antigens. The reverse direction of preexisting antibody responses targeting seasonal hCoVs recognizing SARS-CoV-2 is more difficult to assess. We observed binding of two SARS-CoV-2 epitopes in a few unexposed individuals (peptides from NSP2 in 19% and N protein in 6% (2/32), Table S 2). The abundance of responses against NSP2 did not change in recovered COVID-19 patients, but binding of the N-protein antigen increased to 66% of recovered patients. We also detected additional SARS-CoV-2 peptides bound in single healthy individuals (including a spike peptide, Fig. 2a) similar to a population-wide abundance of 3% (1/32) in unexposed individuals reported by a recent study comparing four antibody tests for the SARS-CoV-2 S/N proteins 28. hCoV peptides bound at higher abundance in unexposed individuals than recovered COVID-19 patients, would suggest a protective nature. As we did not detect any such peptides our data does not support such a simple protective mechanism.

Assessing the protective nature of these population wide preexisting responses would require comparing samples of the same individuals before and after contracting COVID-19 and information on the course (severity) of the disease. The COVID-19 cohort of this study consisted of patients who had experienced mild symptoms, and had not required hospitalization (testing positive in PCR and serological tests). Employing our antigen library to compare antibody profiles against seasonal hCoVs between mild and severe COVID-19 cases could also inform on protective effects of cross-reactivity, as demonstrated for other aspects of the anti-SARS-CoV-2 immune response ^29^ and antibody binding of SARS-CoV-2 proteins ^20^. If severe patients exhibited fewer antibody responses against antigens of seasonal hCoVs, could point towards their protective nature. Moreover, possible detrimental effects of cross-reactive antibody responses originating from seasonal hCoVs leading to antibody-dependent enhancement ^30^ could be assessed with our hCoV antigen library.

### hCov library based high throughput diagnostics

Testing for multiple hCoV antigens at high resolution in parallel could yield higher specificity than conventional tests based on single proteins of SARS-CoV-2 by improving discrimination from seasonal hCoVs. Most current SARS-CoV-2 serological tests rely on detection of entire S-/RBD or N-proteins, reporting an aggregate of binding against all epitopes within ^21,31^. Interpreting the epitope-resolved antibody binding data reported by our assay extends beyond conventional serological tests, as binding against various epitopes of all hCoV needs to be weighed. We used machine learning to build a predictor that highly accurately separated COVID-19 patients from healthy controls based on antibody signatures (AUC=0.96, Fig. 3b). Depending on the intended application and cutoffs employed (Fig. 3c), this assay can display 100% specificity at 72% sensitivity (reporting virtually no false-positives) or 94% specificity at 91% sensitivity. Hence, in addition to informing on cross-reactivity between hCoVs, our antigen library could also represent a tool for SARS-CoV-2 diagnostics.

From a technical perspective, our study shares general limitations of PhIP-Seq, most notable length constraints of presented peptides (64 aa in this study) by underlying oligo synthesis and lack of eukaryotic post translational modifications such as glycosylation. Opposed to neutralization assays carried out with live viruses and cell cultures ^31^, our data does not inform on the neutralizing capacity of the observed binding events. While linear epitopes should be adequately covered, discontinuous, conformational epitopes relying on the correct folding of domains could be missed. We did not frequently detect binding to peptides of the RBD (with one adjacent peptide bound in 25% of COVID-19 and 0% of unexposed individuals’ sera, Fig. 2a), although other work and diagnostic tests relying on the full length RBD had reported common antibody responses in COVID-19 patients ^14,16,17,21^. This discrepancy may be due antibody responses against conformational epitopes in the RBD and/or a lack of S protein glycans ^33^ in the phage displayed peptides.

While current oligo lengths employed in PhIP-Seq may underestimate conformational epitopes, it provides a unique layer of information unobtainable from working with full length antigens or isolated domains: Given the high resolution of the peptide approach, we pinpoint the exact bound regions revealing crucial targets of cross-reactivities. Our hCoV antigen library can be leveraged to study extended cohorts of patients with mild/severe disease courses or samples collected pre/post COVID-19 infection, and could thereby inform on the potential protective nature of preexisting antibody responses against seasonal hCoVs. In addition, our approach can be extended to other antibody isotypes such as IgA, the primary mucosal antibody. Given the low cost of processing phage displayed libraries in parallel ^34^, high accuracy (Fig. 3b,c), and its excellent amenability for robot automation ^12,34^, serological testing based on this hCoV library could be a broadly applicable tool to assess preexisting immunity at population-scale (with implications towards protection and herd immunity of SARS-CoV-2) as well as stratifying vaccine trial costs effectively.

## Data Availability

The majority of data generated or analyzed during this study are included in this published article (and its supplementary information files). Additional datasets generated and analysed during the current study are available from the corresponding author on reasonable request.

## Acknowledgments

E.S.’s COVID-19 research is supported by the Seerave Foundation. T.V. is supported by an Erwin Schrödinger fellowship (J 4256) from the Austrian Science Fund (FWF).

## Materials and methods

### Samples

Serum samples of recovered COVID-19 patients were obtained from MDA (Magen David Adom, the Israeli Red Cross equivalent). These samples had been collected from non-severe cases, who had not been hospitalized. Before sampling, patients had tested twice negative by RT-qPCR testing. Seropositivity of these samples had been confirmed by MDA with a commercial antibody test (Abbot, SARS-CoV-2 IgG, ref. 6R86-22/6R86-32). Control serum samples of unexposed individuals had been collected in 2013/2014 in Israel and reported in a previous study ^25^. Research with the COVID-19 serum samples has been approved by the Weizmann Institute of Science’s institutional review board (#1030-4), and by the Tel Aviv Sourasky Medical Center for the samples of unexposed individuals (#0658-12-TLV). Our cohorts of unexposed individuals and COVID-19 patients showed a different sex distribution and minor age differences (Fig. 1c). While age/sex may influence COVID-19 serology of severe cases ^9,10,38,39^ we do not expect these parameters to affect key conclusions of our study in mildly affected patients (with both cohorts also showing similar antibody responses against viral controls Table S 1b).

### hCoV antigen library design

Reference genomes of the seven hCoVs were downloaded from NCBI directly using amino acid sequences of the translated ORFs with the follow RefSeq accession numbers: SARS-CoV-2 - NC_045512.2, SARS-CoV-1 - NC_004718.3, MERS-CoV - NC_019843.3, hCoV-OC43 - NC_006213.1, hCoV-229E - NC_002645.1, hCoV-HKU1 - NC_006577.2, and hCoV-NL63 - NC_005831.2 For each strain the nonstructural proteins (NSPs) part of the large polyprotein 1ab (polyprotein 1a was discarded if annotated) were separated. The SARS-CoV-2 polyprotein 1ab was cut according to the table published by Wu *et al.^1^*. Additional strains’ polyproteins were processed by the following steps: NSPs 1-3 were cut by sequences reported in the literature ^40^. The remaining NSPs, which are naturally cut by 3Clike protease (3CLpro), were cut by the conserved protease cleavage site (small)-X-(L/I/V/F/M)-Q#(S/A/G), where X is any amino acid and # represents the cleavage position ^41^, and multiply sequence alignment as verification of the site. Specifically for SARS-CoV-2, four additional ORFs reported in the literature ^1^ (but not annotated in RefSeq NC_045512.2) were added.

The final list of proteins were cut to peptides of 64 amino acids (aa) with 20 aa overlaps (to cover all possible epitopes of the maximal length of linear epitope ^42^) between adjacent peptides. The peptide aa sequences were reverse translated to DNA using the *Escherichia coli* codon usage (of highly expressed proteins), aiming to preserve the original codon usage frequencies, excluding restriction sites for cloning (EcoRI and *HindIII*) within the coding sequence (CDS). The coding was re-performed, if needed, so that a barcode was formed in the CDS, by the 44 nt at the 3’ end of each oligo. Every such barcode is a unique sequence at Hamming distance three from all prior sequences in the library, which allows for correcting of a single read error in sequencing the barcode. For similar peptide sequences, alternative codons were used following *E. coli* codon usage, to achieve discrimination. Including the sequencing barcode as part of the CDS, rather than a separate barcode, allowed to use the entire oligo for encoding a peptide (and as opposed to completely omitting a barcode, it did not require sequencing the complete CDS). After finalizing the peptide sequence, the EcoRI and *HindIII* restriction sites, stop codon, and annealing sequences for library amplification were added and obtained from Agilent Technologies as 230mer pool (library amplification primers, fwd: GGACCGCGACTGGAATTCT, rev: CCCGGGCATGAAGCTTTCA) and cloned into T7 phages following the manufacturers recommendations (Merck, T7Select®10-3 Cloning Kit, product number 70550-3).

In this process we had also included controls of viral proteins with high population wide seroprevalence previously reported ^12^ [11 proteins covered by 199 peptides] and negative controls of 42 random peptides and a human protein (SAP4K, 27 peptides) not expected to elicit binding in healthy individuals. A full list of peptides included within the library as well as the corresponding amino acid and nucleotide sequences is provided in supporting file S 3.

### Phage immunoprecipitation sequencing

The PhIP-Seq experiments were performed as outlined in a published protocol ^7^ with the following modifications: PCR plates for the transfer of beads and washing were blocked with 150 µL BSA (30 g/L in DPBS buffer, incubation overnight at 4°C) and BSA was added to diluted phage/buffer mixtures for immunoprecipitations (IPs) to 2 g/L. We reasoned that the Generalized Poisson (GP) distribution approach ^43^ for calling bound peptides may be biased by frequent binding within this hCoV library, leading to a skewed non-binding baseline for estimating GP parameters. Hence, we mixed the hCoV library with a library of 244,000 microbiota and viral antigens that had previously shown a reliable detectability and ratio of bound/unbound peptides (manuscript in preparation). Three microgram of serum IgG antibodies (measured by ELISA) were mixed with the phage library (4,000-fold coverage of phages per library variant). As technical replicates of the same sample were in excellent agreement, measurements were performed in single reactions.

The phage library and antibody mixtures were incubated in 96 deep well plates at 4°C with overhead mixing on a rotator. Forty microliters of a 1:1 mixture of protein A and G magnetic beads (Thermo Fisher Scientific, catalog numbers 10008D and 10009D, washed according to the manufacturers recommendations) were added after overnight incubation and incubated on a rotator for at 4°C. After four hours, the beads were transferred to PCR plates and washed twice as previously reported ^7^ using a Tecan Freedom Evo liquid handling robot with filter tips. The following PCR amplifications for pooled Illumina amplicon sequencing were performed with Q5 polymerase (New England Biolabs, catalog number M0493L) according to the manufacturers recommendations (primer pairs PCR1: tcgtcggcagcgtcagatgtgtataagagacagGTTACTCGAGTGCGGCCGCAAGC and gtctcgtgggctcggagatgtgtataagagacagATGCTCGGGGATCCGAATTC, PCR2: Illumina Nextera combinatorial dual index primers, PCR3 [of PCR2 pools]: AATGATACGGCGACCACCGA and CAAGCAGAAGACGGCATACGA ^7^). PCR3 products were cut from agarose gel and purified twice (1x QIAquick Gel Extraction Kit, 1x QIAquick PCR purification kit; Qiagen catalog numbers 28704/28104) and sequenced on an Illumina NextSeq machine (custom primers for R1: ttactcgagtgcggccgcaagctttca, and for R2: tgtgtataagagacagatgctcggggatccgaattct, R1/R2 44/31 nts). Paired end reads were processed as described below.

### Data analysis

Enriched peptides were calculated (after down-sampling to 1.25 million IDable reads per sample, i.e. reads with a barcode within one error of the set of possible barcodes of the two mixed libraries for which the paired end matched the IDed oligo) by comparing reads of input coverage (library sequencing of phages before IPs) following a Generalized Poisson distribution approach, parameters for which were estimated for each sample separately, as previously reported ^43^. Derived p-values were subject to Bonferroni correction (p-value 0.05) for multiple hypothesis testing, and log-fold-change (number of reads of bound peptides vs. baseline sequencing of phages not undergoing IPs) was computed for all peptides which passed the threshold p-value, all other peptides were given a log-fold-change value of 0.

All oligo creation code, and analysis code was written in Python, using the libraries scikit-learn ^44^, scipy, statsmodels, pandas, numpy and matplotlib.

Alignments shown in Fig. 2 were created with CLC Main Workbench 6 (default settings).Fig. 1

## Supporting information

**S 1:**
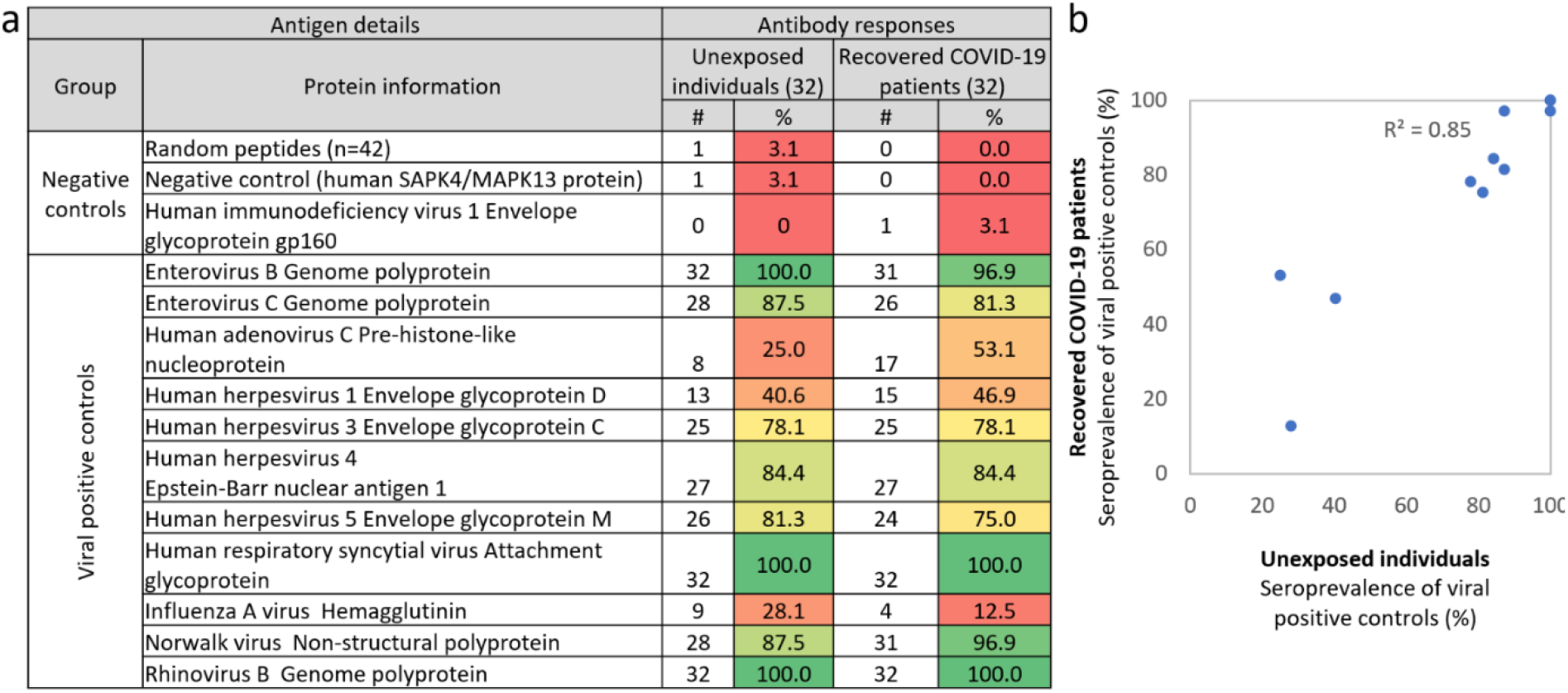
Control antigens included in the library demonstrate low background signal of unspecific binding against random peptides and reproducible antibody recognition of previously reported viral antigens ^12^ with similar abundances between unexposed individuals and recovered COVID-19 patients (b). Peptide negative controls within the hCoV antigen library included 42 random peptides, peptides of the human protein SAPK4 (which should not elicit autoreactive antibodies in healthy individuals), and an HIV protein (with HIV infection being an exclusion criterion for participation in this study). Only 1/42 random peptides and one peptide of the human protein SAPK4 were bound in a single individual, indicating a low background of unspecific binding (or cross-reactivity). Yet, this result indicates that peptides appearing at low signal strengths in single individuals may arise from nonspecific binding. To eliminate such peptides, the following analyses of viral antigens (with population wide seroprevalence previously reported ^12^) was performed using a cut-off of at least two peptides appearing per virus per individual (or peptides occurring in at least four individuals) to count as positive. Following analyses of hCoV binding and predictions were performed with these or even more stringent cutoffs. Serum prevalence for non-hCoV viral antigens were similar between unexposed individuals and recovered COVID-19 patients (b). The slight differences of seroprevalence in unexposed individuals and recovered COVID-19 patients for Human adenovirus C and Influenza A virus antigens may be due the cohort sizes or age/gender differences.

**S 2:**
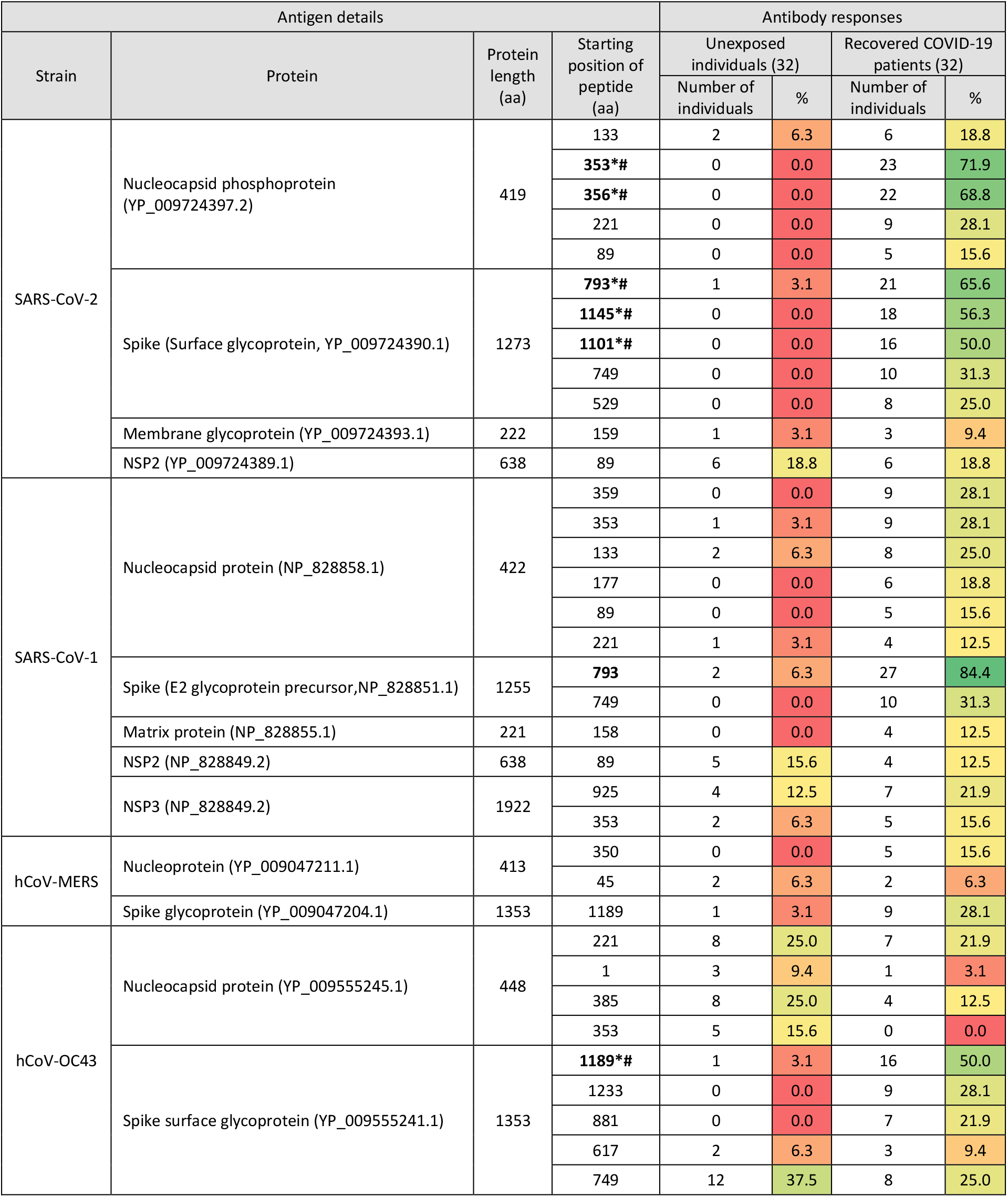
Most frequent hCoV antigens bound in unexposed individuals and recovered COVID-19 patients. Abundance of antibody responses against peptides of hCoV proteins detected in more than four individuals of either cohort are listed. Multiple peptides originating from the same protein are summarized. The seven peptides indicated by bold text are significantly different between unexposed individuals and recovered COVID-19 patients, by scoring the difference between the two distributions of the log fold changes (number of reads of bound peptides vs. baseline sequencing of phages not undergoing IPs) of the two groups. Peptides passing FDR are marked with an ‘*’ and peptides passing also Bonferroni correction with an ‘#’.

**S3:**
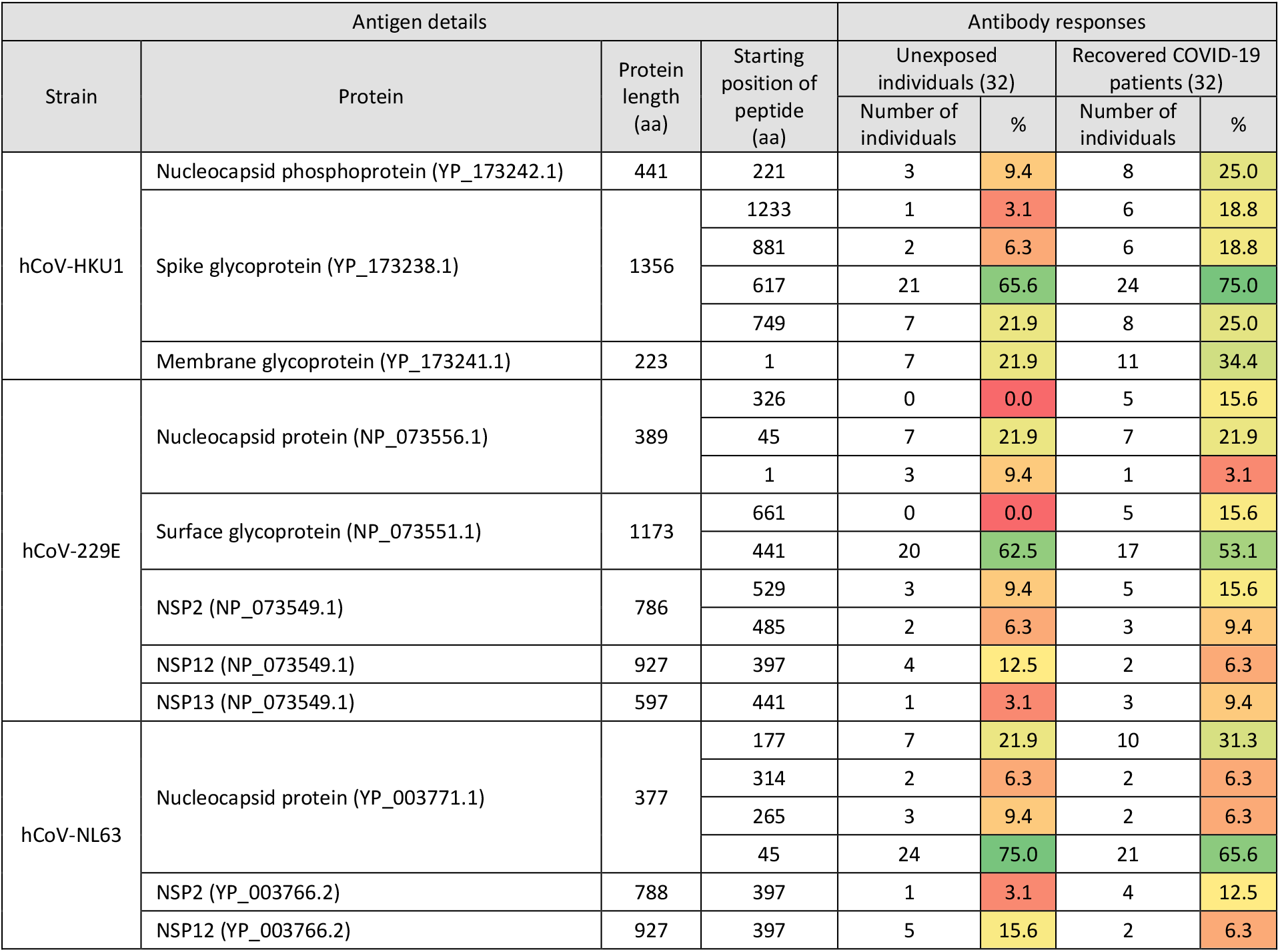
Supporting xslx file with a full list of peptides included within the library as well as the corresponding amino acid and nucleotide sequences (including controls). For each displayed peptide the organism of origin and protein name (including the accession number in the relevant databases NCBI/UniProt/GISAID) is provided. Starting positions of the peptides in the protein’s sequence are provided. Twenty-nine peptides were identical between multiple hCoVs (indicated by the last column ‘Identical to peptide #’) and are highlighted in bold.

## References

1. Wu, A. et al. Genome Composition and Divergence of the Novel Coronavirus (2019-nCoV) Originating in China. Cell Host Microbe 27, 325–328 (2020).

2. Walls, A. C. et al. Structure, Function, and Antigenicity of the SARS-CoV-2 Spike Glycoprotein. Cell 1-12 (2020). doi:10.1016/j.cell.2020.02.058

3. Sette, A. & Crotty, S. Pre-existing immunity to SARS-CoV-2: the knowns and unknowns. Nat. Rev. Immunol. (2020). doi:10.1038/s41577-020-0389-z

4. Grifoni, A. et al. Targets of T Cell Responses to SARS-CoV-2 Coronavirus in Humans with COVID-19 Disease and Unexposed Individuals. Cell 181, 1489-1501.e15 (2020).

5. Le Bert, N. et al. SARS-CoV-2-specific T cell immunity in cases of COVID-19 and SARS. and uninfected controls. Nature (2020). doi:10.1038/s41586-020-2550-z

6. Mateus, J. et al. Selective and cross-reactive SARS-CoV-2 T cell epitopes in unexposed humans. Science (80-.). 21, eabd3871 (2020).

7. Mohan, D. et al. PhIP-Seq characterization of serum antibodies using oligonucleotide-encoded peptidomes. Nat. Protoc. 13, 1958–1978 (2018).

8. Vabret, N. et al. Immunology of COVID-19: current state of the science. Immunity (2020). doi:10.1016/j.immuni.2020.05.002

9. Scully, E. P., Haverfield, J., Ursin, R. L., Tannenbaum, C. & Klein, S. L. Considering how biological sex impacts immune responses and COVID-19 outcomes. Nat. Rev. Immunol. 20, 442–447 (2020).

10. Márquez, E. J., Trowbridge, J., Kuchel, G. A., Banchereau, J. & Ucar, D. The lethal sex gap: COVID-19. Immun. Ageing 17, 1–8 (2020).

11. Woodsworth, D. J., Castellarin, M. & Holt, R. A. Sequence analysis of T-cell repertoires in health and disease. Genome Med. 5, 98 (2013).

12. Xu, G. J. et al. Viral immunology. Comprehensive serological profiling of human populations using a synthetic human virome. Science 348, aaa0698 (2015).

13. Kissler, S. M., Tedijanto, C., Goldstein, E., Grad, Y. H. & Lipsitch, M. Projecting the transmission dynamics of SARS-CoV-2 through the postpandemic period. Science (80-.). 182, eabb5793 (2020).

14. Ju, B. et al. Human neutralizing antibodies elicited by SARS-CoV-2 infection. Nature 584, (2020).

15. Wec, A. Z. et al. Broad neutralization of SARS-related viruses by human monoclonal antibodies. Science (80-.). 736, eabc7424 (2020).

16. Premkumar, L. et al. The receptor binding domain of the viral spike protein is an immunodominant and highly specific target of antibodies in SARS-CoV-2 patients. Sci. Immunol. 5, 1–10 (2020).

17. Yuan, M. et al. A highly conserved cryptic epitope in the receptor-binding domains of SARS-CoV-2 and SARS-CoV. Science 7269, 2020.03.13.991570 (2020).

18. Amanat, F. et al. A serological assay to detect SARS-CoV-2 seroconversion in humans. medRxiv 2020.03.17.20037713 (2020). doi:10.1101/2020.03.17.20037713

19. Kellam, P. & Barclay, W. The dynamics of humoral immune responses following SARS-CoV-2 infection and the potential for reinfection. J. Gen. Virol. (2020). doi:10.1099/jgv.0.001439

20. Atyeo, C. et al. Distinct Early Serological Signatures Track with SARS-CoV-2 Survival. Immunity 1-9 (2020). doi:10.1016/j.immuni.2020.07.020

21. Weissleder, R., Lee, H., Ko, J. & Pittet, M. J. COVID-19 diagnostics in context. Sci. Transl. Med. 12, 1–6 (2020).

22. Barnes, C. O. et al. Structures of human antibodies bound to SARS-CoV-2 spike reveal common epitopes and recurrent features of antibodies. Cell 1-15 (2020). doi:10.1016/j.cell.2020.06.025

23. Jiang, H. wei et al. SARS-CoV-2 proteome microarray for global profiling of COVID-19 specific IgG and IgM responses. Nat. Commun. 11, 1–11 (2020).

24. Li et al., Linear epitope landscape of SARS-CoV-2 Spike protein constructed from 1,051 COVID-19 patients, (2020), medRxiv doi:10.1101/2020.07.13.20152587

25. Zeevi, D. et al. Personalized Nutrition by Prediction of Glycemic Responses. Cell 163, 1079–94 (2015).

26. Gorse, G. J., Patel, G. B., Vitale, J. N. & O’Connor, T. Z. Prevalence of antibodies to four human coronaviruses is lower in nasal secretions than in serum. Clin. Vaccine Immunol. 17, 1875–1880 (2010).

27. World Health Organization, Draft landscape of COVID-19 candidate vaccines - 13 August 2020, https://www.who.int/publications/rn/item/draft-landscape-of-covid-19-candidate-vaccines

28. Grzelak, L. et al. A comparison of four serological assays for detecting anti-SARS-CoV-2 antibodies in human serum samples from different populations. Sci. Transl. Med. 3103, 1–18 (2020).

29. Arunachalam, P. S. et al. Systems biological assessment of immunity to mild versus severe COVID-19 infection in humans. Science 6261, 1–18 (2020).

30. Arvin, A. M. et al. A perspective on potential antibody-dependent enhancement of SARS-CoV-2. Nature 8, 1–11 (2020).

31. Krammer, F. & Simon, V. Serology assays to manage COVID-19. Science (80-.). 368, 1060–1061 (2020).

32. Whitman, J. D. et al. Evaluation of SARS-CoV-2 serology assays reveals a range of test performance. Nat. Biotechnol. (2020). doi:10.1038/s41587-020-0659-0

33. Watanabe, Y., Allen, J. D., Wrapp, D., Mclellan, J. S. & Crispin, M. Site-specific glycan analysis of the SARS-CoV-2 spike. 9983, 1–9 (2020).

34. Larman, H. B. et al. PhIP-Seq characterization of autoantibodies from patients with multiple sclerosis, type 1 diabetes and rheumatoid arthritis. J. Autoimmun. 43, 1–9 (2013).

35. Wrapp, D. et al. Cryo-EM structure of the 2019-nCoV spike in the prefusion conformation. Science 367, 1260–1263 (2020).

36. Cubuk et al. (2020) The SARS-CoV-2 nucleocapsid protein is dynamic, disordered, and phase separates with RNA. preprint, bioRxiv doi:10.1101/2020.06.17.158121

37. Chen, T. & Guestrin, C. XGBoost: A Scalable Tree Boosting System. in Proceedings of the 22nd ACM SIGKDD International Conference on Knowledge Discovery and Data Mining 785-794 (ACM, 2016). doi:10.1145/2939672.2939785

38. Takahashi, T. et al. Sex differences in immune responses that underlie COVID-19 disease outcomes. Nature (2020). doi:10.1038/s41586-020-2700-3

39. Bunders, M. & Altfeld, M. Implications of sex differences in immunity for SARS-CoV-2 pathogenesis and design of therapeutic interventions. Immunity 2, 1–9 (2020).

40. Yang, X. et al. Proteolytic processing, deubiquitinase and interferon antagonist activities of Middle East respiratory syndrome coronavirus papain-like protease. J. Gen. Virol. 95, 614–626 (2014).

41. Snijder, E. J., Decroly, E. & Ziebuhr, J. The Nonstructural Proteins Directing Coronavirus RNA Synthesis and Processing. Adv. Virus Res. 96, 59–126 (2016).

42. Forsströ, B. et al. Dissecting antibodies with regards to linear and conformational epitopes. PLoS One 10, 1–11 (2015).

43. Larman, H. B. et al. Autoantigen discovery with a synthetic human peptidome. Nat. Biotechnol. 29, 535–41 (2011).

44. Buitinck et al. API design for machine learning software: experiences from the scikit-learn, (2013) ECML PKDD Workshop: Languages for Data Mining and Machine Learning 108-122

